# Towards early risk biomarkers: serum metabolic signature in childhood predicts cardio-metabolic risk in adulthood

**DOI:** 10.1101/2019.12.11.19014308

**Authors:** Xiaowei Ojanen, Runtan Cheng, Timo Törmäkangas, Na Wu, Noa Rappaport, Tomasz Wilmanski, Wei Yan, Nathan D. Price, Sulin Cheng, Petri Wiklund

## Abstract

Cardiovascular diseases have their origin in childhood. Early biomarkers identifying individuals with increased risk for disease are needed to support early detection and to optimize prevention strategies. By applying machine learning approach on high throughput NMR-based metabolomics data, we identified metabolic predictors of cardiovascular risk in circulation in a cohort of 396 females, followed from childhood (mean age 11.2 years) to early adulthood (mean age 18.1 years). The identified childhood metabolic signature included three circulating biomarkers robustly associating with increased cardiovascular risk in early adulthood (AUC = 0.641 to 0.802, all p<0.01). These associations were confirmed in two validation cohorts including middle-aged women, with similar effect estimates. We subsequently applied random intercept cross-lagged panel model analysis, which suggested causal relationship between metabolites and cardio-metabolic risk score from childhood to early adulthood. These results provide evidence for the utility of circulating metabolomics panel to identify children and adolescents at risk for cardiovascular disease, to whom preventive measures and follow-up could be indicated.

## Background

Cardiovascular diseases are the largest contributors to global mortality and morbidity^1^, and a significant economic burden to the healthcare system^2^. Cardio-metabolic abnormalities, including hyperglycemia, elevated blood pressure, dyslipidemia and abdominal obesity are risk factors for the development of cardiovascular diseases^3-6^. Although the clinical complications of cardiovascular disease typically manifest in adulthood, autopsy and observational studies have shown that development of atherosclerosis starts in childhood and adolescence, and is associated with the same cardiovascular disease risk factors that are well established in adults^7^. Early identification of children who are at risk of developing cardiovascular disease would allow instituting and maintaining optimum health behaviors, at a time when it is likely to be most effective.

The availability of metabolic screens provides an opportunity to identify biomarkers associated with cardiovascular risk. For example, using liquid chromatography/mass spectrometry Cheng et al. demonstrated that obesity, hypertension, insulin resistance and dyslipidemia were associated with multiple circulating metabolites including branched-chain and aromatic amino acids in adults^8^. These same metabolites have also been consistently associated with future development of type 2 diabetes^9^ and cardiovascular disease^10^. Metabolomics profiling studies in children and adolescents, however, have reported conflicting results; most of these previous studies are cross-sectional, and the few existing longitudinal studies have short follow-up durations and small number of participants 11-17. Therefore, longitudinal studies examining temporal associations between the circulating metabolome and cardio-metabolic risk factors from childhood to adulthood are needed. In this study, we used an NMR-based metabolomics platform to quantify 121 circulating metabolic measures in children followed longitudinally from pre-puberty to early adulthood. Our results show that a small pre-pubertal metabolic signature predicts cardio-metabolic risk score in adulthood, and provides evidence for a causal role of atherogenic lipoprotein particles and systemic low-grade inflammation in cardiovascular disease pathogenesis.

## Results

### Cardio-metabolic risk score

We first examined cardio-metabolic risk factor clustering, i.e. metabolic syndrome, defined as the presence of at least three of the following five risk factors: abdominal obesity (waist circumference >88 cm), elevated blood pressure (≥130/85 mmHg), elevated fasting blood glucose (≥5.6 mmol/L), elevated serum triglycerides (≥1.7 mmol/L) and reduced serum high-density lipoprotein cholesterol (HDL-C) (<1.29 mmol/L)^18^ in a total of 396 Finnish adolescent females that participated in a longitudinal study, from pre-puberty (mean age, 11.2 years) to early adulthood (mean age, 18.1 years) (**Table 1**). At the 7.5 years follow-up, 3.2% of the participants were classified as having metabolic syndrome according to this definition. In addition, 10.8% of the participants had two risk factors, 40.1% had one risk factor, and 45.9% had no positive cardio-metabolic risk factors. Due to the low prevalence of metabolic syndrome and relatively high prevalence of two and single risk factors in this cohort, we constructed a continuously distributed variable to represent clustering of cardio-metabolic risk factors referred to as “MetS” score from here on^19^. The MetS score was calculated similarly to previously published scores^20-22^ by standardizing and then summing (separately for each measurement wave) the following continuously distributed metabolic traits: mean arterial pressure ([(2 x diastolic blood pressure) + systolic blood pressure]/3); abdominal fat mass; fasting plasma glucose; serum high-density lipoprotein cholesterol^-1^; and fasting serum triglyceride z-scores. A higher score indicates a higher cardio-metabolic risk.

**Table 1.** General characteristics at different measurement time points in adolescent girls

| Variables | Baseline<br>(n=215) |  | 2-year follow-up<br>(n=192) |  | 7.5-year follow-up<br>(n=222) |  |
| --- | --- | --- | --- | --- | --- | --- |
|  | Mean | 95% CI | Mean | 95% CI | Mean | 95% CI |
| Age (years) | 11.2 | 11.1 – 11.5 | 13.3 | 13.0 – 13.5 | 18.1 | 17.8 – 18.4 |
| Height (cm) | 146.3 <sup>a,b</sup> | 145.0 – 147.6 | 158.0 <sup>c</sup> | 156.9 – 159.1 | 165.1 | 163.7 – 166.5 |
| Weight (kg) | 39.7 <sup>a,b</sup> | 38.4 – 41.1 | 50.1 <sup>c</sup> | 48.5 – 51.6 | 60.3 | 59.0 – 61.6 |
| BMI (kg/m<sup>2</sup>) | 18.4 <sup>a,b</sup> | 18.0 – 18.9 | 20.0 <sup>c</sup> | 19.5 – 20.5 | 21.9 | 21.5 – 22.3 |
| Lean mass<sub>WB</sub> (kg) | 26.5 <sup>a,b</sup> | 26.0 – 27.1 | 33.9 <sup>c</sup> | 33.2 – 34.5 | 38.2 | 37.6 – 38.7 |
| Fat mass<sub>WB</sub> (kg) | 10.7 <sup>a,b</sup> | 10.0 – 11.4 | 13.8 <sup>c</sup> | 12.8 – 14.8 | 19.5 | 18.4 – 20.7 |
| Fat mass<sub>Abdominal</sub> (kg) | 0.789 <sup>a,b</sup> | 0.72 – 0.86 | 1.09 <sup>c</sup> | 0.98 – 1.19 | 1.45 | 1.35 – 1.55 |
| Systolic BP (mmHg) | 106 <sup>a,b</sup> | 104 – 107 | 111 <sup>c</sup> | 109 – 113 | 119 | 117 – 120 |
| Diastolic BP (mmHg) | 62 <sup>b</sup> | 61 – 63 | 64 <sup>c</sup> | 63 – 66 | 71 | 70 – 72 |
| IGF1 (mmol/l) | 31.7 <sup>a</sup> | 30.1 – 33.2 | 47.5 <sup>c</sup> | 46.0 – 49.1 | 32.3 | 31.3 – 33.4 |
| E2 (nmol/L) | 0.116 <sup>b</sup> | 0.11 – 0.13 | 0.187 <sup>c</sup> | 0.17 – 0.20 | 0.376 | 0.29 – 0.46 |
| Te (nmol/L) | 0.779 <sup>a,b</sup> | 0.67 – 0.89 | 1.52 <sup>c</sup> | 1.37 – 1.66 | 4.10 | 3.44 – 4.76 |
| SHBG (nmol/L) | 82.6 <sup>a,b</sup> | 78.3 – 86.8 | 64.3 <sup>c</sup> | 60.6 – 67.9 | 119 | 104 – 133 |
| Leptin (ng/mL) | 20.8 | 18.4 – 23.1 | 23.1 | 20.7 – 25.5 | 22.8 | 20.7 – 25.0 |
| Adiponectin (µg/mL) | 11594 | 10833 – 12355 | 11293 | 10600 – 11986 | 11731 | 10830 – 12632 |
| 25(OH)D (nmol/L) | 45.9 <sup>a</sup> | 43.9 – 47.9 | 42.0 <sup>c</sup> | 39.8 – 44.2 | 46.2 | 44.2 – 48.2 |
| PTH (pg/mL) | 25.9 <sup>a</sup> | 24.6 – 27.1 | 28.8 <sup>c</sup> | 27.4 – 30.3 | 24.0 | 22.5 – 25.5 |
| Glucose (mmol/L) | 5.50 <sup>b</sup> | 5.44 – 5.57 | 5.41 <sup>c</sup> | 5.35 – 5.48 | 5.27 | 5.19 – 5.34 |
| Insulin (IU/mL) | 8.71 <sup>a</sup> | 8.24 – 9.81 | 12.47 <sup>c</sup> | 11.3 – 13.7 | 8.37 | 7.77 – 8.97 |
| HDL (mmol/L) | 1.42 | 1.39 – 1.46 | 1.46 | 1.42 – 1.50 | 1.47 | 1.43 – 1.51 |
| Triglycerides (mmol/L) | 0.878 <sup>b</sup> | 0.836 – 0.919 | 0.951 | 0.897 – 1.01 | 1.01 | 0.962 – 1.06 |
| Energy intake (kcal/day) | 1564 <sup>a,b</sup> | 1517 – 1610 | 1739 | 1675 – 1803 | 1780 | 1717 – 1843 |
| Protein (E%) | 15.5 <sup>b</sup> | 15.1 – 16.0 | 15.5 <sup>b</sup> | 15.0 – 16.1 | 17.3 | 16.8 – 17.8 |
| Fat (E%) | 33.5 <sup>b</sup> | 32.7 – 34.2 | 32.5 | 31.7 – 33.4 | 31.7 | 30.7 – 32.7 |
| Carbohydrates (E%) | 51.0 | 50.0 – 51.9 | 51.9 | 50.9 – 53.0 | 50.2 | 49.1 – 51.3 |
| LTPA (score) | 149 | 130 – 168 | 166 | 145 – 186 | 174 | 148 – 201 |
| IPA (Hours/day) | 18.3 <sup>b</sup> | 18.1 – 18.6 | 18.2 | 17.9 – 18.6 | 17.7 | 17.3 – 18.1 |

Data are given as mean and their 95 % CI. Natural logarithm transformation data were used for the comparison of different time points. The Sidak method was used for multiple comparisons. BMI = body mass index (weight (kg)/height (m)<sup>2</sup>), E% = percentage of daily energy intake, LTPA = leisure-time physical activity, IPA = sitting + lying down, IGF1 = Insulin-like growth factor 1, E2 = Estradiol, Te = Testosterone, SHBG = sex hormone binding globulin, 25(OH)D = 25-hydroxyvitamin D, PTH = intact parathyroid hormone, HDL = high-density lipoprotein cholesterol.
<sup>a</sup> p<0.05 baseline compared with 2-year follow-up, <sup>b</sup> p<0.05 baseline compared with 7-year follow-up, <sup>c</sup> p<0.05 2-year compared with 7.5-year follow-up

### Correlation network of circulating metabolites with cardio-metabolic risk score

We next explored cross-sectional associations of serum metabolites (**Supplementary Figure 1**) with the MetS score at each measurement wave (baseline, 2-year and 7.5-year follow-up) to assess the stability of associations during the course of pubertal growth. Several lipoprotein subclasses, high-density lipoprotein diameter (HDL-D), HDL-2C and VLDL-TG, apolipoprotein B to apolipoprotein A1 ratio (ApoB/ApoA1) and glycoprotein acetyls (GlycA) were consistently associated with MetS score at all three time points after correcting for multiple hypothesis testing (**Figure 1**, inner cycle **and Supplementary Figure 1a and b and Supplementary Table 2**). In addition, we found associations of apolipoprotein A1 and omega 6 fatty acids with the MetS score at baseline and 2-year follow-up, very-low-density lipoprotein (VLDL-D) at baseline and 7.5-year follow-up, fatty acid length and triglycerides/phosphoglycerides ratio (TG/PG) at 2-year follow-up, and apolipoprotein B isoleucine, monounsaturated fatty acids (MUFA) and TG/PG at 7.5-year follow-up. Further adjustment for physical activity, dietary intake and other covariates (i.e., sex hormones, SHBG and IGF-1, insulin, adiponectin, leptin and vitamin D) did not materially change the results. The significant correlations between metabolites and MetS score are presented in **Supplementary Table 3**.

**Figure 1.**
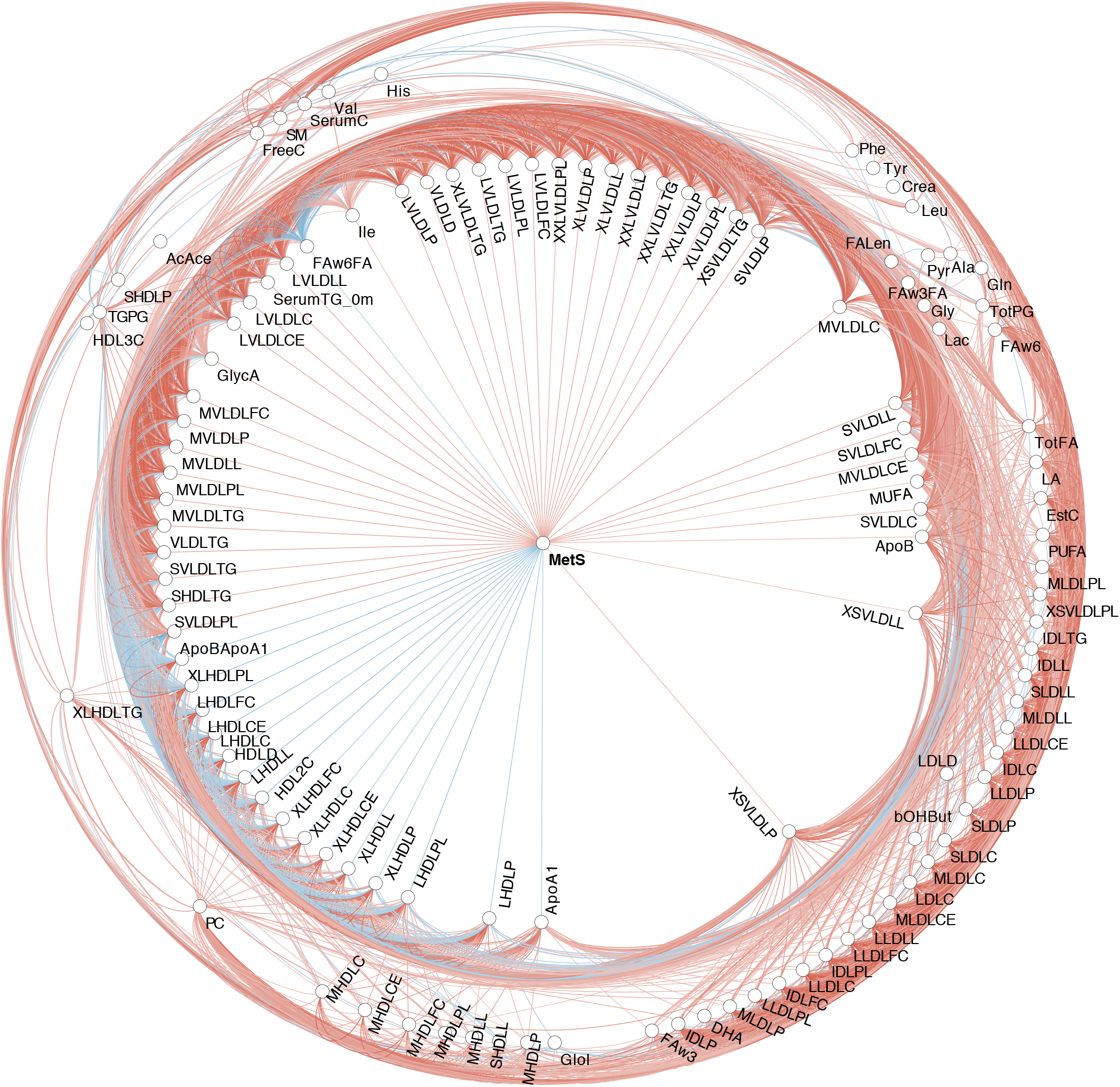
Bivariate Spearman correlation network among metabolites and between metabolites and MetS at baseline. Each node represents a variable while the edges linking variable pairs represent significant correlations between the corresponding nodes. Significant correlations between metabolites and MetS are shown in the inner cycle. There are 118 nodes with 2483 edges.

### Serum metabolites predict future MetS score

We used the Least Absolute Shrinkage and Selection Operator (LASSO) method to identify circulating metabolites that predict the MetS score from pre-puberty to early adulthood using a five-fold cross-validation scheme (CV) (**Supplementary File 1)**. Of the 121 metabolites measured for each study participant, ten metabolites at baseline and 11 metabolites at 2-year follow-up were retained in the final model. These subsets of metabolites were predictive of MetS score at 7.5 year follow-up, explaining on average 20.1% (r^2^ ranged from −0.39 to 0.55) and 11.9% (r^2^ ranged from −0.26 to 0.63) of the variance, respectively (**Figure 2a,b,d,e**, **Supplementary File 1 and Supplementary Table 4**). The identified metabolites, included different amino acids, glycolysis and inflammation related metabolites, ketone bodies, fatty acids, apolipoproteins, and lipoprotein subclass particles. Four metabolites (glutamine, L-HDL-PL, ApoB/ApoA1 ratio, GlycA) were retained in the model at both time points, showing greater associations with increasing age. **(Supplementary Table 4**).

**Figure 2.**
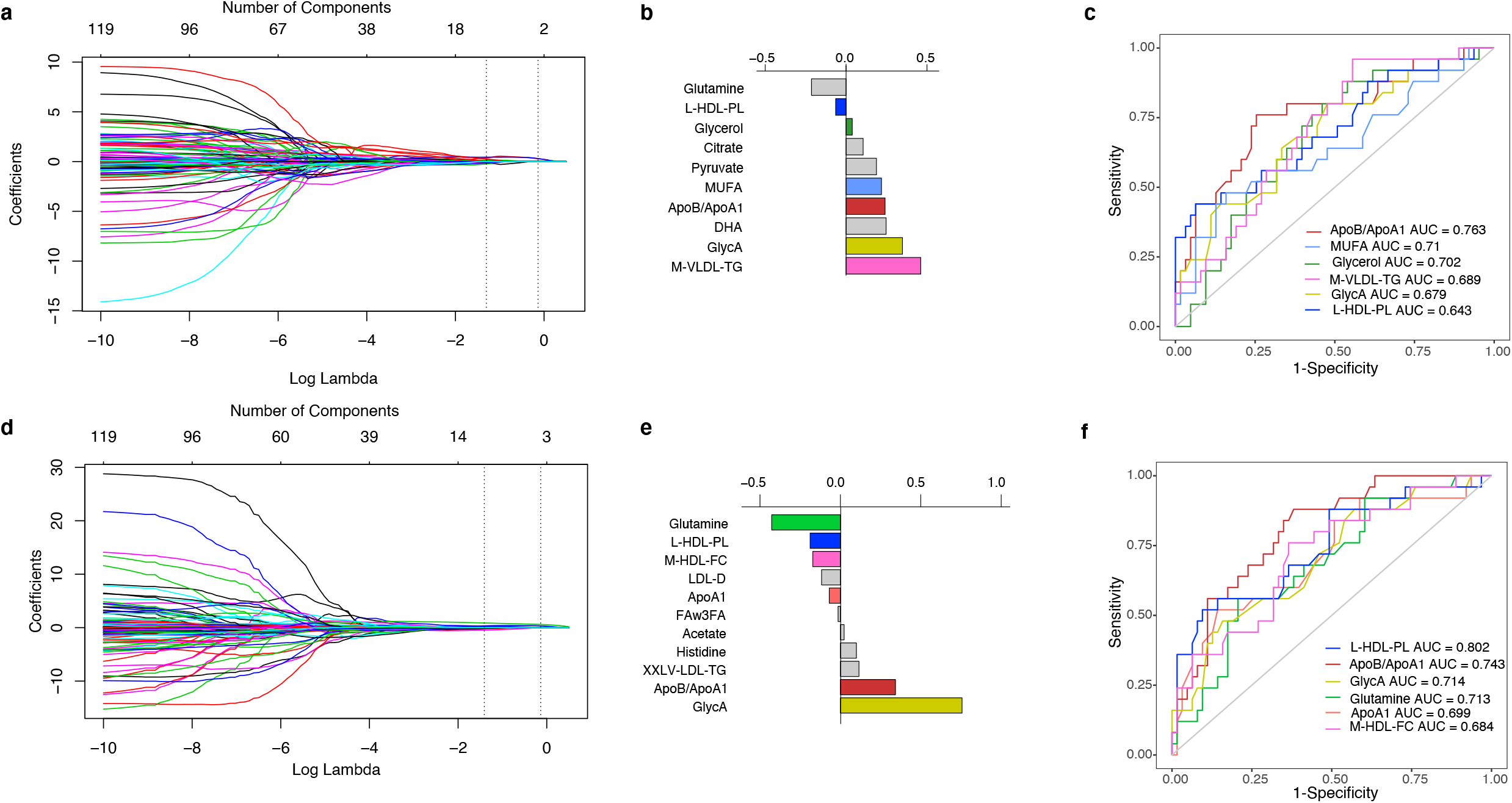
LASSO regression identifies metabolite predictors of adult MetS score. The upper panel represents metabolites at baseline predicted MetS score at 7.5-year follow-up. a and d: The vertical dashed lines indicate the LASSO fit for which the mean squared error is smallest. The lower x-axis represents LASSO parameter lambda while the upper x-axis represents the number of variables with non-zero coefficients. The y-axis represents LASSO regression coefficient which is depending on the value of lambda. b and e: LASSO regression coefficients of selected metabolites. c and f: The area under the curve of these significant metabolites (AUC) by Receiver Operating Characteristics (ROC) analysis. ApoB/ApoA1 = apolipoprotein B to apolipoprotein A1 ratio; GlycA = glycoprotein acetyls; HDL2C = high-density lipoprotein two cholesterol; M-VLDL-TG = medium lipoprotein triglycerides; L-HDL-PL = very large high-density lipoprotein phospholipids; M-HDL-FC = medium high-density lipoprotein free cholesterol; XXLV-LDL-TG = extremely large very-low-density lipoprotein triglycerides; His = Histidine.

### Performance of cardio-metabolic risk prediction

To test the ability of the metabolites to distinguish individuals with high cardio-metabolic risk from those with low risk we stratified the study participants into quartiles based on their MetS score values at 7.5 year follow-up and performed Receiver Operating Characteristics (ROC) analysis. The area under the curve (AUC) indicated that of the metabolites identified by LASSO at baseline, ApoB/ApoA1, MUFA, Glycerol, GlycA and L-M-VLDL-TG and L-HDL-PL were able to classify individuals with high MetS score (AUC: 0.643 to 0.763, p = 0.001 to p < 0.0001, Figure 2c), while L-HDL-PL ApoB/ApoA1,GlycA, Glutamine, ApoA1,and M-HDL-FC were significant classifiers at the 2-year follow-up (AUC: 0.684 to 0.802, p = 0.03 to p < 0.0001, **Figure 2f**).

### Directional influences between the serum metabolites and cardio-metabolic risk

To assess the direction of causality between the metabolites and MetS score associations, we applied Random Intercept Cross-lagged panel analysis model (RI-CLPM, **Figure 3a**), which is a multilevel structural equation model that partitions the between-person variance from the within-person variance, thus allowing estimating cross-lagged effects both at between-person and at within-person level, while controlling for correlations within time-points and autoregressive effects, or stability, across time. The autoregressive part of the model showed that the previous MetS score predicted the subsequent MetS score at each time point from childhood to early adulthood. Similar effects were observed between the repeated measurements of ApoB/ApoA1 and GlycA and L-HDL-PL (all p < 0.001, **Figure 3b and c**) Likelihood ratio tests indicated a significant between-subjects effect in the model when ApoB/ApoA1 (χ2df_=3_ = 17.0, p = 0.001, **Figure 3b**), GlycA (χ2df_=3_ = 10.2, p = 0.017, **Figure 3c**), or L-HDL-PL (χ2df_=3_ =12.1, p = 0.007, **Figure 3d**), were considered, while the within-subject effects did not have a significant contribution for either ApoB/ApoA1 (χ2df_=4_ = 3.7, p = 0.442) or GlycA (χ2df_=4_ = 4.6, p = 0.333) or L-HDL-PL (χ2df_=4_ = 8.6, p = 0.072).

**Figure 3.**
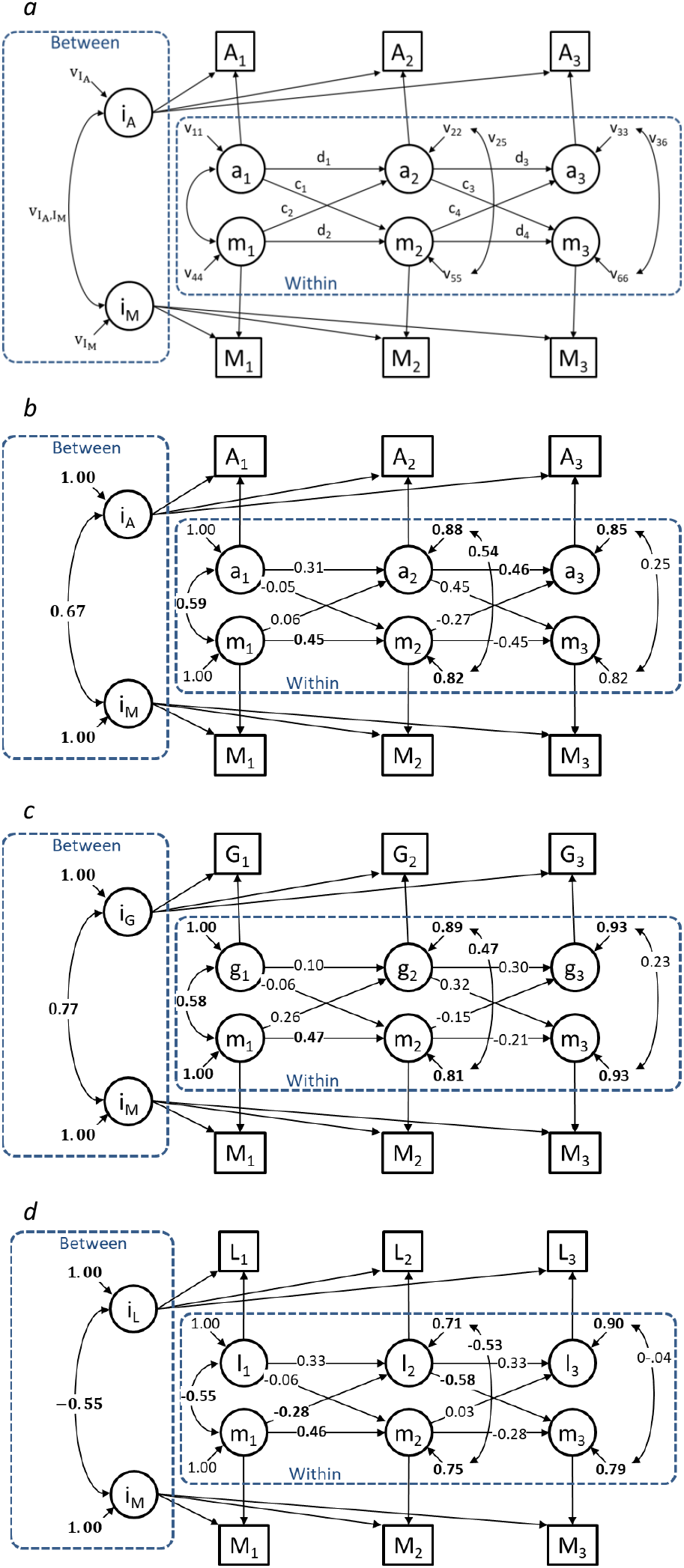
Directional influences between the common metabolite predictors. a: Conceptual random intercept cross-lagged panel model (RI-CPLM). The path coefficients of main interest include the between-subjects random intercept variances 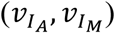 and covariance 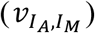 and within-subjects autoregressive coefficients (d) and cross-lagged coefficients (c). The remaining parameters (v) include variances / residual variances and correlations / residual correlations among the variables. Directed arrows with no coefficient attached were constrained to unity. b: The RI-CPLM for ApoB/ApoA1; c: the RI-CPLM for GlycA; d: the RI-CPLM for L-HDL-PL.

Causal predominance was examined by comparing standardized coefficients of the cross-lagged paths, and the results suggested a causal effect of baseline ApoB/ApoA1 on MetS score at 2-year follow-up, while MetS score at 2-year follow-up had causal effect on ApoB/ApoA1 at 7.5-year follow-up (**Figure 3b**). Similarly, the MetS score at baseline had a causal effect on L-HDL-PL at 2-year follow-up, while L-HDL-PL at 2 year-follow-up had a causal effect on MetS score at 7.5 year follow-up (**Figure 3d**). The results also suggested causal predominance of MetS score at baseline and at 2 year-follow-up on GlycA both at 2-year and 7.5-year follow-ups, but associations were not statistically significant (**Figure 3c**). All RI-CLPM results are shown in **Supplementary Table 5**.

### Key metabolite predictors are confirmed in two validation cohorts

To validate our findings, we explored the main results in two other datasets. The first included the discovery cohort participants’ sisters (n = 74) and their biological mothers (n = 206). The other included a cohort of middle-aged overweight and obese pre-menopausal women (n = 105). Characteristics of the participants are shown in **Supplementary Table 6**. Among the sisters, 5.2% of the participants had metabolic syndrome, 16.9% had two positive risk factors, 40.3% had one positive risk factor, and 37.7% had no positive cardio-metabolic risk factors. In mothers, 12.6% of the participants had metabolic syndrome, 20.2% had two positive risk factors, 28.2% had one positive risk factor, and 20.2% had no positive cardio-metabolic risk factors. In pre-menopausal women, 26.4% of the participants had metabolic syndrome, 36.8% had two positive risk factors, 31.1% had one positive risk factor, and 5.7% had no positive cardio-metabolic risk factors.

As the validation cohort includes overweight and obese women with more cardio-metabolic risk factors, this allowed us to compare the concentrations of the metabolite biomarkers identified in the discovery cohort between healthy women and women with metabolic syndrome, to validate that the differences observed in puberty persist into adulthood (**Figure 4a and Supplementary Table 7**). Furthermore, in mothers (**Figure 4b and Supplementary Table 7**) and sisters (**Figure 4c and Supplementary Table 7**), we compared the high risk and low risk groups by quartiles (the highest quartile represents high risk).

**Figure 4.**
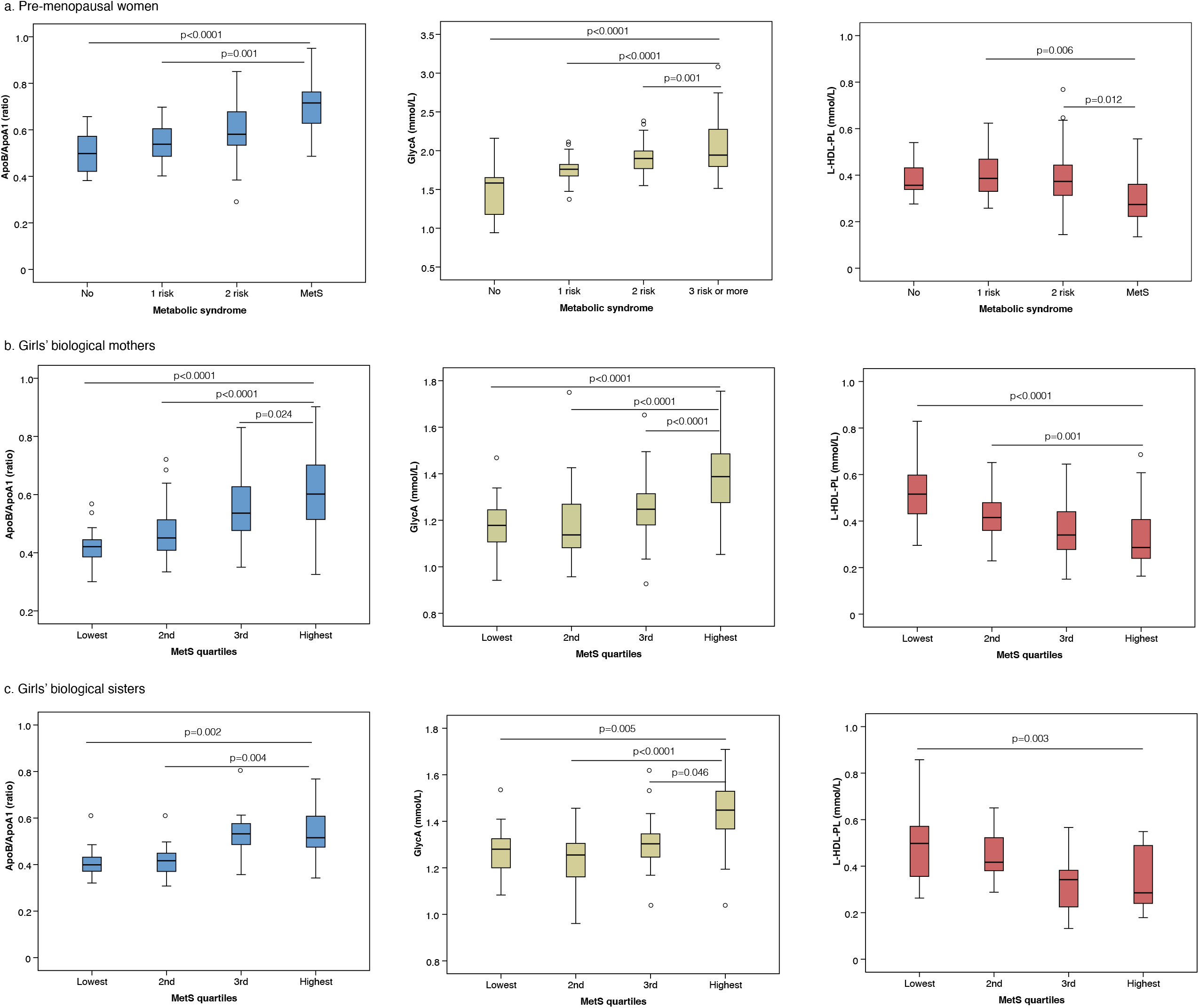
Comparison of different cardio-metabolic risk groups in validation cohorts for selected metabolites. a: premenopausal women by comparison of metabolic syndrome (MS) to 1-risk factor, 2-risk factors and without MS (NMS); b: biological mothers by comparison of MetS quartiles; c: biological sisters by comparison of MetS quartiles. Plot boxes represent median with 95% confident interval (CI). The circles out of the plot boxes are the outliers corresponding to each group.

We further used LASSO for these two validation data sets and the performance was similar to the discovery dataset (**Supplementary file 2)**. In the sisters and mothers, the explained variance of metabolites predicting MetS was similar to pre-menopausal women (r^2^ = 0.51 and r^2^ = 0.54, respectively, **Supplementary file 2**) which were comparable to the discovery cohort at baseline (r^2^ = 0.77) and 7.5-year follow-up (r^2^ = 0.58). The AUCs were similar to the discovery cohort as well (**Figure 5 a, b and c and Supplementary Table 8**). This analysis validates that the identified metabolites are elevated in women who are at high risk of cardio-metabolic disease.

**Figure 5.**
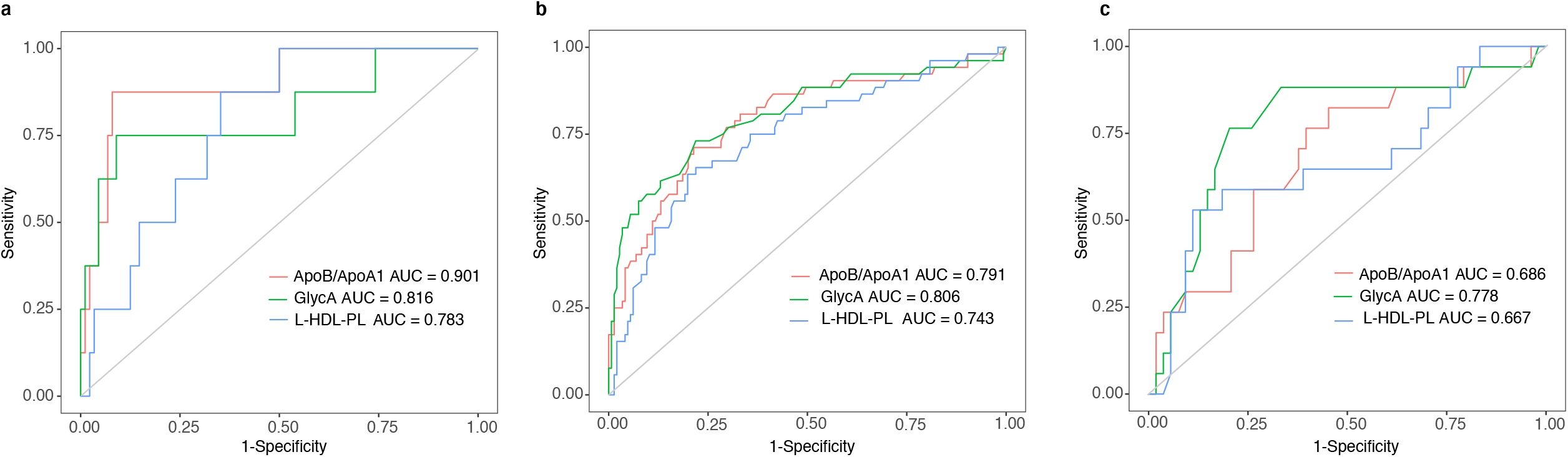
Validation for performance of the selected metabolites retained by LASSO to predict cardio-metabolic risk. a: premenopausal women; b: biological mothers; c: biological sisters. The area under the curve of these significant metabolites (AUC) by Receiver Operating Characteristics (ROC) analysis. ApoB/ApoA1 = apolipoprotein B to apolipoprotein A1 ratio; GlycA = glycoprotein acetyls; L-HDL-PL = very large high-density lipoprotein phospholipids.

## Discussion

In this study, we utilized a rich longitudinal data resource to identify a metabolic signature in childhood that predicts increased cardiovascular disease risk in adulthood. These metabolic measures have atherogenic properties and reflect chronic systemic inflammation, previously associated with future cardiovascular disease and pre-mature mortality in older individuals. Thus, our results provide further evidence that the pathogenic processes that contribute to cardiovascular diseases in later life originate in childhood and adolescence, providing an impetus to earlier intervention strategies to reduce the global burden of cardiovascular disease.

We found that the serum ApoB/ApoA1 ratio in childhood strongly and consistently predicted future cardio-metabolic risk score (MetS) across all time points. ApoB and ApoA1 are the two main lipoproteins involved in lipid transport. ApoB is the main protein in VLDL and LDL (atherogenic) particles, while ApoA1 is the main protein in HDL (anti-atherogenic) particles. This result is supported by an earlier 9-year follow-up study in school girls from childhood to adulthood, demonstrating that ApoB/ApoA1 ratio was associated with metabolic syndrome and it’s components^23^. In another prospective study, Juonala et al. reported that ApoB and ApoA1 levels and their ratio in adolescence were associated with carotid artery intima-media thickness and brachial artery flow-mediated dilation in adulthood^24^. In that study, ApoB and ApoA1 were stronger predictors of abnormal vascular changes than conventional cholesterol measurements (LDL-C and HDL-C), which suggests that the carriers (apolipoproteins) might play a more central role than the actual lipid content transported in these lipoprotein particles. These findings are further supported by the Pathobiological Determinants of Atherosclerosis in Youth study which found apolipoproteins associated with postmortem arterial lesions^25^, and the Bogalusa Heart Study, which showed that high ApoB/ApoA1 ratio in children was associated with incidence of parental myocardial infarction^26^.

We also found strong inverse association between L-HDL phospholipids in childhood and cardio-metabolic risk in early adulthood. A previous study by Piperi et al. found that HDL-phospholipids were more closely related to coronary artery disease than HDL-C or other lipoproteins studied ^27^, and Meikle et al. recently reported that HDL phospholipids, but not HDL cholesterol, distinguished acute coronary syndrome from stable coronary artery disease^28^. Low serum HDL-phospholipid concentrations have also been associated with high coronary artery calcification scores in asymptomatic patients with atherosclerosis^29^, and with increased risk of metabolic syndrome and coronary heart disease, particularly in women^30^. Our key finding of the strong inverse association between L-HDL phospholipids in childhood and cardio-metabolic risk in early adulthood was confirmed in both validation cohorts, demonstrating that adult women with metabolic syndrome had lower serum L-HDL-PL concentration than their healthy counterparts. It has been previously shown that HDL phospholipids play an important role in the cholesterol efflux process^31^, and that metabolic syndrome is associated with progressive reduction in cholesterol efflux capacity, which contribute to development of atherosclerosis32. Taken together, our results suggest that exposure to an atherogenic apolipoprotein profile and low HDL phospholipids in childhood may cause reduced cholesterol efflux capacity and changes in the arteries that contribute to the development of atherosclerosis and coronary heart disease in adulthood.

It is increasingly recognized that the atherosclerotic process involves not only lipid and lipoprotein metabolism but it also requires a pro-inflammatory response that includes both the innate and acquired immune systems ^33^. Another metabolite predicting MetS score in our study was GlycA, which is a novel marker of systemic inflammation reflecting signals originating from glycan groups of acute-phase glycoproteins, mainly a1-acid glycoprotein, but also other acute-phase reactants such as haptoglobin, a1-antitrypsin, a1-antichymotrypsin, and transferrin^34^. GlycA has been found to correlate with adiposity, insulin resistance and other markers of metabolic syndrome in adults ^35, 36^, suggesting that in addition to being elevated in acute inflammation, GlycA might also be biomarker of subclinical vascular inflammation ^37^. Accordingly, it was recently shown that plasma GlycA is independently associated with the incidence of cardiovascular disease in a large cohort study of initially healthy women^37^. Elevated circulating GlycA levels have also been shown to predict risk of type 2 diabetes^38^, nonalcoholic fatty liver disease^39^, and all-cause mortality^40^ in adults, but so far, no comparable data are available in children and adolescents. In our study, GlycA levels in childhood and adolescence strongly and consistently predicted MetS score in early adulthood, and this finding was confirmed in both validation cohorts. Thus, the findings our study confirm and extend the results of earlier reports by demonstrating that GlycA is viable biomarker of systemic subclinical inflammation associated with increased cardio-metabolic risk not only in adults but also in children and adolescents. However, further studies are needed before GlycA could be adopted as a routinely used in clinical test for the purposes of risk assessment, management, or follow-up in pediatric populations.

We further investigated the direction of associations between metabolite biomarkers and cardio-metabolic risk in a random intercept cross-lagged path model. The results revealed a consistent pattern in which increased ApoB/ApoA1 ratio in childhood was associated with an increased MetS score in adolescence, and MetS score in adolescence was associated with increased ApoB/ApoA1 ratio in early adulthood. Similar sequence of reciprocal cause and effect was observed between the MetS score and the other two key metabolites i.e. L-HDL-PL and GlycA from childhood to early adulthood, suggesting that metabolites and cardiovascular risk factors aggravate each other, leading inexorably to a worsening cardio-metabolic health.

The major strength of our study is the longitudinal data on females followed from pre-puberty to early adulthood, which enabled us to explore the temporal associations between circulating metabolites and cardio-metabolic risk during pubertal growth. We were also able to validate the main results in females of different ages in three other datasets. Furthermore, downstream analyses allowed us to determine the specificity and sensitivity of the most important metabolite biomarkers, and explore the direction of causation between metabolites and MetS. However, some limitations warrant consideration. The cohort included only females which limits the generalization of the results. Furthermore, the prevalence of metabolic syndrome in adolescents was low. However, while binary definition of metabolic syndrome might be a useful tool for clinical practice to assess cardiovascular disease risk in adults^18^, continuous MetS score is more appropriate for epidemiological studies for the following reasons: first, dichotomizing continuous outcome variables reduces statistical power^41^; second, the risk of cardiovascular disease is an aggregative progressive function of several risk factors^42^, and third, cardiovascular risk increases progressively with increasing numbers of risk factors^43^. Therefore, multiple studies with pediatric and adult populations have used continuous metabolic risk scores, integrating components of the metabolic syndrome definition to represent clustering of metabolic risk factors.

## Conclusions

We identified a robust signature of serum metabolites, including ApoB/ApoA1 ratio, GlycA, and L-HDL-PL, which may serve as early biomarkers for increased cardio-metabolic risk in children and adolescents. Further studies are needed to explore the generalizability and the potential clinical utility of these biomarkers to lower the global burden of cardiometabolic disease through shaping early-life health strategies.

## Methods

### Study setting

The study was conducted in the city of Jyväskylä, and the surrounding area in Central Finland. Circulating metabolite concentrations were analyzed using a high-throughput serum nuclear magnetic resonance (NMR) metabolomics platform that quantified 121 metabolite measures including routine lipids, lipoprotein subclass distributions, particle size and composition, fatty acids and other low-molecular-weight metabolites such as amino acids and glycolysis-related metabolites (all metabolites measures in the platform are shown in **Supplementary Table 1**). In addition, behavioral characteristics, including leisure-time physical activity, dietary intake as well as medical history were collected via validated self-administered questionnaires. Body composition was assessed using dual-energy X-ray absorptiometry (DXA). Sex-hormones, sex-hormones binding globulin (SHBG), insulin-like growth factor 1 (IGF-1), insulin, leptin, adiponectin, parathyroid hormone and 25-hydroxyvitamin D were determined using standard protocols.

### Subjects

A total of 396 girls (mean age 11.2 years at baseline) participated in a longitudinal study at different time points for an average of 7.5 years. Detailed meta-data regarding the participants and study design have been reported previously^44, 45^. Briefly, the subjects were first contacted via class teachers of grades 4 to 6 (age 9 to 13 years old) in 61 schools in the city of Jyväskylä and its surroundings, located Central Finland (96% of all the schools in these areas). For this report, we included subjects with valid data on body composition, cardio-metabolic risk markers and serum metabolomics. Thirteen individuals who reported using oral contraceptives at the age of 18 were excluded from this study. The final discovery cohort dataset included 215 girls at baseline (100% were pre-menarche), 192 at 2-year (38% were pre-menarche) and at 222 7.5-year follow-up (100% post-menarche) (**Table 1**).

Cross-sectional data from the discovery cohort participant’s sisters (aged 18.6 years, n = 74) and their biological mothers (aged 48.4 years, n = 206) and an independent cohort of Finnish pre-menopausal women (aged 41.5 years, n = 105) was included in the study for the purpose of validation of results obtained from the discovery cohort (**Supplementary Table 6**).

Written informed consent was obtained from all participants and their parents. The study was conducted in accordance with the Declaration of Helsinki and approved by the Ethic Committee of the Central Hospital of Central Finland and the Finnish National Agency of Medicines (memo 22/8/2008 and 5/2009).

### Background information

Health history and lifestyle characteristics were collected via self-administered questionnaires at the time of laboratory tests (baseline, 24- and 7.5-year follow-up). Dietary intake of total energy and energy-yielding nutrients was assessed from three-day food records (2 week days and 1 weekend day) using Micro-Nutrica software (Social Insurance Institution, Turku, Finland) as described elsewhere^46^). Leisure time physical activity (PA), including walking, jogging, running, gym fitness, ball games, swimming, etc., expressed as hours/week and times/week, and a score of PA was evaluated using a validated self-administered PA questionnaire, as previously described^47^.

### Anthropometry and body composition assessments

Body height was measured using a stadiometer. Body weight was measured using an electronic scale with subjects wearing only underwear. BMI was calculated by dividing body weight in kilograms by the square of the body height (in meters). Blood pressure (BP) in the right arm was recorded using an automated oscillometric method (OMRON M3 Intellisense, OMRON Healthcare, Co., Ltd, Kyoto, Japan) in a sitting position in the morning after a 10-minute rest. Two consecutive measurements were performed, and the mean of the measurements was used. Menarche age was de?ned as the ?rst onset of menstrual bleeding and was collected by questionnaire, retrospective phone call, and/or interview during a clinical visit.

Lean tissue mass, and fat mass of the whole body, and abdominal region were assessed using DXA (Prodigy GE Lunar Corp., Madison, WI USA). Precision of the repeated measurements expressed as a coefficient of variation (CV%) were 1.0% for LM, and 2.2% for FM.

### Circulating biomarker assessments

Blood samples from the antecubital vein were collected between 7 and 9 AM after an overnight fast. The samples were collected during early follicular phase (2 to 5 days after the beginning of menstruation) in subjects with regular menses. All samples were handled and analyzed by a qualified phlebotomist using slandered protocols. Serum and plasma were extracted from blood by centrifugation and stored immediately at −80°C until analyzed. Plasma glucose was assessed by KONELAB 20XTi analyzer (Thermo Fischer Scientific inc. Waltham, MA, USA). Insulin was measured by immunofluorescence using the IMMULITE Analyser (Diagnostic Products Corporation, Los Angeles). The inter- and intra-assay CVs were 2.0% and 3.7 % for glucose and 11% and 3.4% for insulin, respectively.

Serum leptin was assessed using human leptin (ELISA; Diagnostic Systems Laboratories, Inc., Webster, TX) and adiponectin was measured by an enzymeimmunoassay method using the Quantikine Human Total Adiponectin/Acrp30 Immunoassay (R&D Systems). Insulin-like growth factor 1 (IGF-1) was assessed using time-resolved fluoroimmunoassays (IMMULITE; Siemens Healthcare Diagnostics, IL, USA). Estradiol (E2), testosterone (Te) and sex hormone binding globulin (SHBG) were determined using ELISA (NovaTec Immundiagnostica GmbH, Dietzenbach, Germany). 25-hydroxyvitamin D (25(OH)D) was measured by radioimmunoassay (Incstar Corporation, Stillwater, MN) and intact parathyroid hormone (PTH) concentrations was measured by using an immunoradiometric method (Nichols Institute, Juan San Capistrano, CA). Inter- and intra-assay coefficients of variation (CVs) were 4.6%, 2.7% and 2.2% for leptin, 3.3% and 4.3% for adiponectin, 6.1% and 3.1% for IGF-1, 3.2% and 5.4% for E2, 3.9%, 6.2% for Te, and 1.1% and 1.1% for SHBG, 10% and 15% for 25(OH)D, and 4% and 3% for PTH, respectively.

Serum metabolite concentrations were analyzed using a high-throughput nuclear magnetic resonance (NMR) metabolomics platform. The experimental protocols, including sample preparation and NMR spectroscopy, have been described in detail elsewhere^48^. This methodology combines three molecular windows that contain the majority of the metabolic information available by ^1^H NMR from native serum such as serum lipids, cholesterol, lipoprotein subclasses as well as various low-molecular-weight metabolites, including amino acids, ketone bodies and glycolysis intermediates. The final number of metabolites included in this report was 121 (full list of metabolites measured in this study are shown in **Supplementary Table 1**).

### Cardio-metabolic risk assessment

To assess cardio-metabolic risk, a standardized continuously distributed variable for clustered metabolic risk (MetS score) was constructed similar to previously published scores ^20-22^. The risk score was calculated by standardizing and then summing the following continuously distributed metabolic traits: mean arterial pressure ([(2 x diastolic blood pressure) + systolic blood pressure]/3); abdominal fat mass; fasting plasma glucose; serum HDL cholesterol x −1; and fasting serum triglyceride z-score. The z-scores for each variable and MetS scores were calculated separately for each time point. A higher score indicates a higher cardio-metabolic risk.

### Statistical analysis

Continuous data were tested for normality by Shapiro-Wilk’s test before each analysis. If data were not normally distributed, their natural logarithms were used in all analyses. Descriptive statistics are presented as means and 95% confidence interval (CI).

Correlation network: Bivariate Spearman correlation between metabolites (n=121) and MetS was used to calculate correlation coefficients for each time point by using R statistics and the correlation network was drawn using Cytoscape version 3.7.1. In the network, each node represents a variable while the edges represent significant correlations between the connected nodes. Multiple testing correction between MetS and metabolites as well as among metabolites was performed using the Bonferroni method with an adjusted p-value cutoff of 0.05. The metabolites that were significantly correlated with MetS after Bonferroni correction are presented in **Supplementary Table 2**.

In addition, Bivariate Spearman partial correlation was performed controlling for hormones (insulin, leptin, adiponectin, IGF-1, E2, Te, SHBG, 25(OHD) and PTH, PA score, and energy (Kcal/day) and energy yield micronutrients (protein, carbohydrate and fats E%).

LASSO regression: To predict future cardio-metabolic risk, we applied LASSO analysis using the R GLMNET package^40^. The parameters of the LASSO used in this report are given in supplemental material (**Supplementary File 1**). In the LASSO analysis, MetS from baseline to 2-year and 7.5 year follow-up was treated as a continuous variable and only girls who had data for both time points were included (baseline to 2-year paired n=172, baseline to 7.5 year paired n=88 and 2-year to 7.5-year paired n=89). We performed a 5-fold cross-validation. In each fold, 80% of the data was used to train the LASSO model and then performance was evaluated using the 20% of the data not used for model optimization. This process was repeated five times on each of the five folds of the data, resulting in 5 test-set out-of-sample r-square values of five different models. This method obtain a conservative estimate of model performance (**Supplementary File 1**).

### Diagnostic performance of cardio-metabolic risk

To assess the “diagnostic performance” i.e. specificity and sensitivity of the LASSO-identified high cardio-metabolic risk by metabolites, we stratified the study participants into quartiles based on their MetS score values. The highest quartile was considered as the high-risk group. For premenopausal women, the high-risk group was defined by women with metabolic syndrome (MS). We then performed AUROC (Area Under the Receiver Operating Characteristics) analysis using R plot ROC package. The area under the curve (AUC) was reported in **Figure 2c and f** only for those metabolites which were significant with 95% confidence interval (CI).

Cross-lagged paths model (CLPM): The CPLM is based on an empirically-motivated definition of causality^49^, where the association is considered to have a causal interpretation, if the preceding measurement of a predictor explains the current response while adjusting for the preceding measurement of the response. The parameters of main interest include directed paths for the auto-regressive relationships, which can be viewed as measures of the temporal stability of the variables across time-points, and cross-lagged relationships, which are used to assess predictive relationships between the variables at consecutive time points. Because the cross-lagged effects are represented as path coefficients and not correlations, it is possible that the estimate of the effect from the predictor to the response can take an opposite sign when compared to the observed correlation between those variables. This difference is due to the fact that the predictor’s effect is partialled out from the previous measurement of the response. The bidirectional associations between the identified metabolites and MetS were analyzed by Mplus version 7.4.

Validation: We validated the performance of the biomarkers retained by LASSO using R (v.3.6.0) GLMNET (v.2.0) package in two other cross-sectional data sets (participants’ sisters and their biological mothers and in a cohort of unrelated pre-menopausal women). 121 metabolites overlapping with the ones measured in the discovery cohort were used as input, and the prediction of MetS was output. R squared value of the correlation between predicted and observed MetS was calculated to represent the model performance. All training and test data were scaled and standardized to mean 0 and standard deviation 1 before analysis (**Supplementary File 2**).

In addition, we compared the highest quartile to the lowest 3 quartiles in sisters and mothers, and NMS to 1-risk, 2-risk and MetS in premenopausal women by analysis of variance followed by Sidak for multiple group comparison (**Figure 4 and Supplementary Table 7**). Then, we compared those with MetS to those without MetS by ROC (**Figure 5 and Supplementary Table 8**) as described in above diagnostic performance of cardio-metabolic risk section.

## Data Availability

Data will available upon request

## Data availability

Availability of the data will follow the data management principles for research at the University of Jyväskylä https://www.jyu.fi/tutkimus/tutkimusaineistot/rdmenpdf. Established researchers wishing to collaborate will be given access to the de-identified data following approval of a signed research proposal.

## Acknowledgements

Funding agencies: This study was financially supported by the Academy of Finland, Ministry of Education of Finland and University of Jyväskylä. The National Nature Science Foundation of China (Grant 31571219), the 111 Project (B17029), the Shanghai Jiao Tong University Zhiyuan Foundation (Grant CP2014013), and China Postdoc Scholarship Council (201806230001).

Disclosure: The authors declared no conflict of interest.

## Author information

XO, PW and SC participated in data collection, data analysis and drafted the manuscript. XO, RT, NW, WY, TT performed the data analysis. RT, NW, NR, TW, WY, TT and NDP edited the manuscript. PW, SC, NR, TW and NDP designed the study. All authors approved the submitted version. PW and SC had full access to all of the data in the study and take responsibility for the integrity of the data and the accuracy of the data analysis.

## Supplementary information

The supplementary files include data processing scripts, Supplementary Figures and Tables.

**Supplementary File 1**: LASSO regression identifies metabolite predictors of subsequent MetS score data processing scripts.

**Supplementary File 2**: LASSO regression identifies metabolite predictors of MetS score data processing script in validation cohorts.

**Supplementary Figure 1**. Bivariate Spearman correlation network among metabolites and between metabolites and MetS. Each node represents a variable, while the edges linking variable pairs represent significant correlations between the corresponding nodes. Significant correlation between metabolites and MetS are shown in the inner cycle. a: 2-year follow-up, there are 117 nodes with 2422 edges; b: 7.5-year follow-up, there are 122 nodes with 3412 edges.

**Supplementary Table 1**. Metabolites list used in this report.

**Supplementary Table 2**. Bivariate Spearman correlation coefficient of significant metabolites with MetS in Figure 1a, b and c after Bonferroni correction for multiple tests.

**Supplementary Table 3**. Partial correlations between metabolites and MetS controlled for hormones, physical activity and dietary intake.

**Supplementary Table 4**. Lasso regression coefficient of significant predictors for MetS.

**Supplementary Table 5**. Random intercept cross-lagged panel analysis for potential reciprocal relationship between metabolites and MetS score.

**Supplementary Table 6**. Validation datasets of participants’ biological sisters and their biological mothers and unrelated pre-menopausal women.

**Supplementary Table 7**. Comparison of selected metabolites by metabolic risk groups.

**Supplementary Table 8**. Validation of the “diagnostic performance” using ROC analysis.

## Tables

**Table 1.** Subjects’ background.

## Notes

### Competing Interest Statement

The authors have declared no competing interest.

